# Intraoperative localization and preservation of reading in ventral occipitotemporal cortex

**DOI:** 10.1101/2021.11.11.21266202

**Authors:** Oscar Woolnough, Kathryn M. Snyder, Cale W. Morse, Meredith J. McCarty, Samden D. Lhatoo, Nitin Tandon

## Abstract

Resective surgery in language-dominant ventral occipitotemporal cortex (vOTC) carries the risk of causing impairment to reading. As it is not on the lateral surface, it is not easily accessible for intraoperative mapping and extensive stimulation mapping can be time consuming. Here we assess the feasibility of using task-based electrocorticography (ECoG) recordings intraoperatively to help guide stimulation mapping of reading in vOTC.

In 11 patients undergoing extraoperative, intracranial seizure mapping we recorded induced broadband gamma activation (70 – 150 Hz) during a visual category localizer. Word-responsive cortex localized in this manner showed a high sensitivity (72%) to stimulation-induced reading deficits, and the confluence of ECoG and stimulation positive sites appears to demarcate the visual word form area.

In two additional patients, with pathologies necessitating resections in language-dominant vOTC, task-based functional mapping was performed intraoperatively using subdural ECoG, alongside direct cortical stimulation. Cortical areas critical for reading were mapped and successfully preserved, while enabling pathological tissue to be completely removed. Data collection is possible in <3 minutes and initial intraoperative data analysis takes <3 minutes, allowing for rapid assessment of broad areas of cortex.

Eloquent cortex in ventral visual cortex can be rapidly mapped intraoperatively using ECoG. This method acts to guide high-probability targets for stimulation, with limited patient participation, and can be used to avoid iatrogenic dyslexia following surgery.

## Introduction

Ventral occipitotemporal cortex (vOTC) contains several cortical regions dedicated to higher-level visual processing of scenes, faces, objects and words, that follow a well-characterized, medial-to-lateral topography with regards to their preferred tuning.^1–3^ Lesions of any of these category selective regions can result in isolated deficits in a categorical domain, such as prosopagnosia or scene agnosia. ^4,5^ In particular, the visual word form area (VWFA), located in the lateral aspect of language-dominant vOTC, displays category selectivity to written words, as demonstrated by functional imaging and direct cortical stimulation,^6–9^ and lesions that result in alexia.^10,11^ Given the critical role of reading in modern life, it is imperative to localize and preserve reading sites in vOTC to enable safe resective procedures in this region.

Direct cortical stimulation during an awake craniotomy is the widely accepted gold-standard for localizing and preserving critical language regions.^12,13^ This enables optimal resections while preserving cognitive function.^14,15^ However, performing intraoperative stimulation of the ventral surface of the temporo-occipital region is challenging given limited access, additional intraoperative time, and a high level of patient engagement required for comprehensive mapping of reading function.^13^ Electrocorticography (ECoG) has been widely used in the extraoperative setting to localize seizure foci in patients with drug-resistant epilepsy and for basic insight into cognitive functions. Previous studies have shown that extraoperative ECoG and cortical stimulation can co-localize language processes,^16–19^ including for naming,^20^ speech,^21^ and reading^8,9^ tasks, in reliable and concordant fashion. Furthermore, mapping category selectivity of word-selective cortex with these tasks can be performed rapidly and does not necessitate subject participation.^22^ This suggests that task-based ECoG may be an effective method of guiding and increasing efficiency of intraoperative stimulation of vOTC. Here we assess the use of extraoperative and intraoperative recordings alongside stimulation mapping to identify and preserve eloquent reading cortex in lateral vOTC while enabling resection of pathological tissue.

## Materials and Methods

### Participants

13 patients (6 male, 18-47 years, all right-handed, IQ 94 ± 8, Age of Epilepsy Onset 18 ± 9 years) were semi-chronically implanted with intracranial electrodes for seizure localization of pharmaco-resistant epilepsy. Eleven patients participated in an extraoperative visual category localizer task and underwent extraoperative cortical stimulation mapping. Two patients (P1, P2), detailed below, performed an intraoperative category localizer and underwent direct cortical stimulation during an awake resective craniotomy. All participants gave written informed consent and all experimental procedures were reviewed and approved by the Committee for the Protection of Human Subjects (CPHS) of the University of Texas Health Science Center at Houston as Protocol Number HSC-MS-06-0385.

### Functional Imaging

Task-based fMRI data were acquired using a gradient-recalled echo-planar imaging sequence consisting of 33 axial slices with 3 mm thickness and in-plane resolution of 2.75 mm isotropic (TE = 30 ms, TR = 2,015 ms, flip angle = 90°).

### Category Localizer

Stimuli were presented on an LCD screen positioned at eye-level, approximately 50 cm from the patient. 60 (P1, Extraoperative) or 40 (P2) each of (i) words, (ii) faces, (iii) consonant strings and (iv) phase-scrambled faces were presented in pseudorandom order at an approximate size of 9° visual angle (Figure 1A).^22^ Stimuli were presented using Psychophysics Toolbox in MATLAB. Each image was displayed for 750 ms (P1, Extraoperative) or 500 ms (P2) with an inter-stimulus interval of 500 ms. Total stimulus presentation time for P1 was <7 minutes and for P2 was <3 minutes. To ensure attention, 20 images were presented twice in a row, which the patients indicated by either pressing a button (Extraoperative) or saying “Repeat” aloud (P1, P2). Repeat trials were not included in the analysis.

**Figure 1:**
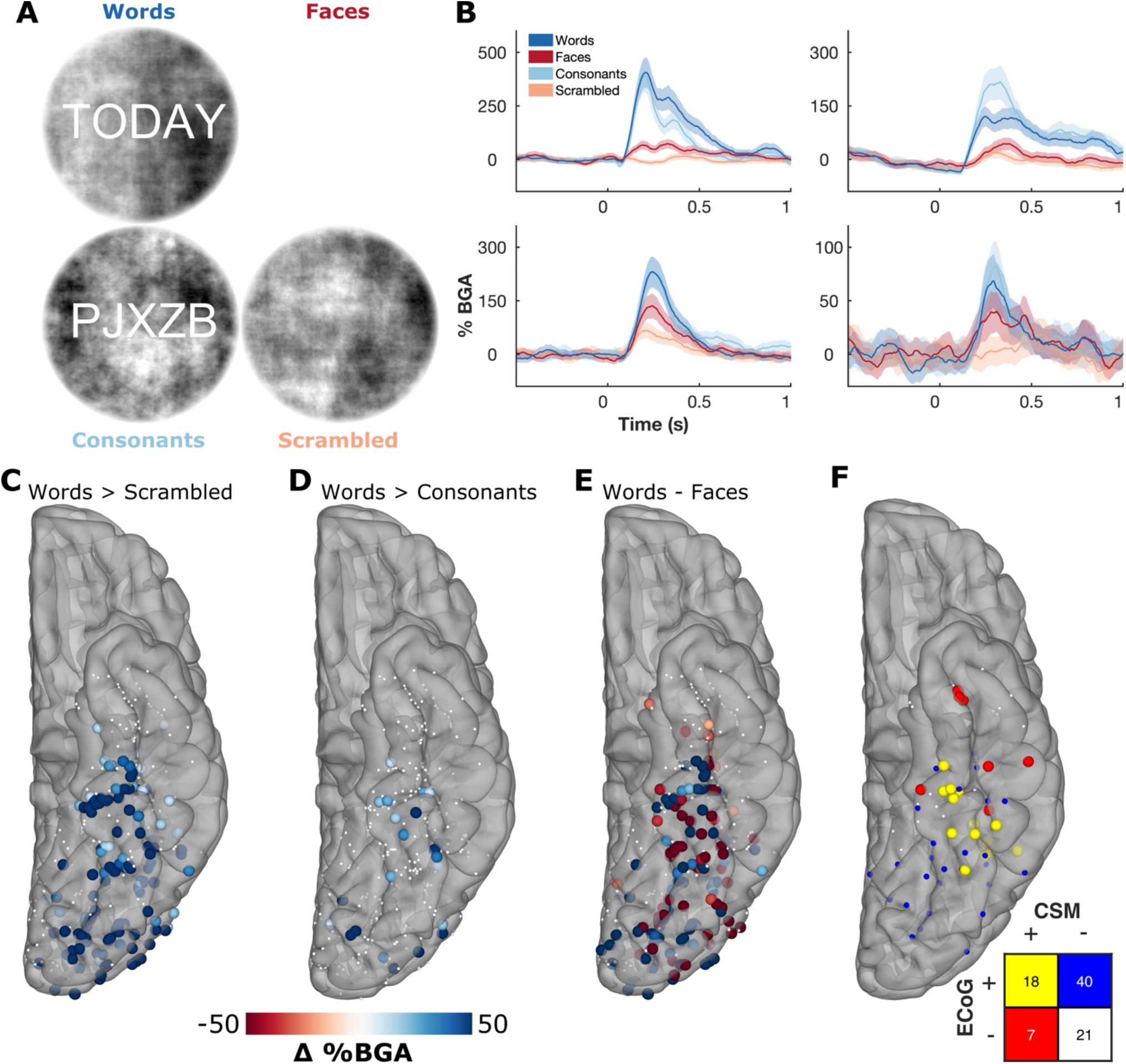
Extraoperative ECoG Category Selectivity. A: Example stimuli from the visual category localizer. B: Time course of broadband gamma for representative stimulation-positive electrodes from four patients. C-E: Contrasts of words > scrambled (C), words > consonants (D), and words – faces (E) in the window 100-400 ms after stimulus onset. Thresholded by electrodes with a significant difference (two-sample t-test, p<0.01). F: Concordance between task-related activation (words > scrambled; ECoG+) and cortical stimulation mapping (CSM) sites, in ventral occipitotemporal cortex, leading to reading disruption (CSM+). CSM was performed between neighboring electrode pairs. Pairs were considered ECoG+ if either electrode was significant.

### Signal Analysis

Extraoperative data were acquired from either subdural grid electrodes (2 patients) or stereotactically placed depth electrodes (9 patients). Subdural electrodes were platinum-iridium electrodes embedded in a silicone elastomer sheet (PMT Corporation; top-hat design; 3mm diameter cortical contact), and were surgically implanted via a craniotomy.^23,24^ Stereotactic probes were implanted using a Robotic Surgical Assistant (Medtech, Montpellier, France).^25,26^ Each probe (PMT corporation, Chanhassen, Minnesota) was 0.8 mm in diameter and had 8-16 electrode contacts. Each contact was a platinum-iridium cylinder, 2.0 mm in length and separated from the adjacent contact by 1.5 - 2.43 mm.

For the intraoperative cases, ECoG electrodes were subdural platinum-iridium electrodes embedded in a silicone elastomer sheet (PMT Corporation, Manhasset, MN; top-hat design; 3mm diameter cortical contact, 0.66 cm interelectrode spacing). Electrodes were temporarily secured in place by suturing the connection leads to the dural edges.^23,24,27^ Data were digitized at 2 kHz using the NeuroPort recording system (Blackrock Microsystems, Salt Lake City, Utah).

In the extraoperative cases, signals were re-referenced to the common average of the clean channels while for the intra-operative cases signals were referenced to an electrode on the lateral cortical surface, away from visually response regions. Trials contaminated by inter-ictal epileptic spikes were discarded. Broadband gamma activity (BGA; 70-150Hz) was extracted with a frequency domain bandpass Hilbert transform (paired sigmoid flanks with half-width 1.5 Hz) and the analytic amplitude was smoothed (Savitzky - Golay finite impulse response, 3^rd^ order, frame length of 201 ms). BGA is presented here as percentage change from baseline level, defined as the period −500 to −100 ms before stimulus presentation. Initial data analysis and visualization for intraoperative cases was performed in the operating room, on a laptop running a MATLAB-based pipeline, in <3 minutes. Extraoperative electrode localization was performed by co-localizing a pre-operative MRI with the post-implantation CT.^27^ For post-surgical visualization of intraoperative cases, electrodes were localized manually in AFNI, based on intraoperative photographs and localizations using a surgical navigation system (StealthStation S8, Medtronic). Cortical surface reconstruction was performed using FreeSurfer and imported into AFNI where the electrode positions were mapped onto the cortical surface.^27,28^

### Cortical Stimulation Mapping

Cortical stimulation mapping (CSM) was carried out using a Nihon Kohden PE-210A stimulator through the ECoG electrodes (Extraoperative), or with an OCS2 handheld stimulator (5mm electrode spacing; Integra LifeSciences, France) (P1, P2). 50Hz, 500 µs square pulse stimulation was administered with a current between 5 and 10 mA depending on an established baseline that did not result in after-discharges.^23^ Testing included reading of standardized passages (i.e. the Grandfather, Rainbow and North Wind passages).^9^ Intraoperatively, visual stimuli were presented on the same screen used for the ECoG task stimuli and auditory stimuli were delivered orally by the surgeon.

## Results

### Extraoperative Reading Mapping

We used three different ECoG contrasts to predict stimulation induced reading deficits: (i) words > scrambled (72% sensitivity), (ii) words > consonants (24% sensitivity) and (iii) words > faces (56% sensitivity) (Figure 1C-E). The words > scrambled contrast showed the greatest overlap with the CSM-positive sites and the region of greatest concordance between ECoG and CSM was located within mid-fusiform cortex - the predicted location of the VWFA (Figure 1F).

Within mid-fusiform cortex, discordant results between ECoG and CSM commonly occurred at electrodes neighboring true positive electrodes. This likely reflects the lower resolution of stimulation as mostly non-overlapping pairs of electrodes were stimulated. Patients with CSM-positive, ECoG-negative electrodes in the anterior temporal lobe were notably present in patients with epileptic foci in proximity to the site of stimulation and reported dizziness, confusion and aura-like symptoms during stimulation of these sites.

### Intraoperative Category Localizer

Two patients scheduled to undergo awake resective craniotomies in the language-dominant vOTC were recruited for intraoperative task-based ECoG mapping in addition to clinical standard language mapping.

<u>Patient 1 (P1)</u> was a teenage right-handed female with medication-resistant focal epilepsy, having failed treatment with Lacosamide and Lamotrigine. Seizures started as a loss of awareness with subsequent asymmetric tonic limb posturing, a right sided figure of 4 sign and right-sided facial twitching. This evolved into right hemibody clonus and then a generalized tonic-clonic seizure. Her MRI revealed a lesion in the left inferior temporal gyrus consistent with a focal cortical dysplasia (FCD) (Figure 2A-D). EEG showed inter-ictal spikes and seizure onset localized to the left posterior temporal lobe. Clinical onset of seizures preceded electrographic manifestations on EEG. Magnetoencephalographic source reconstruction localized inter-ictal discharges in the vicinity of the FCD.

**Figure 2:**
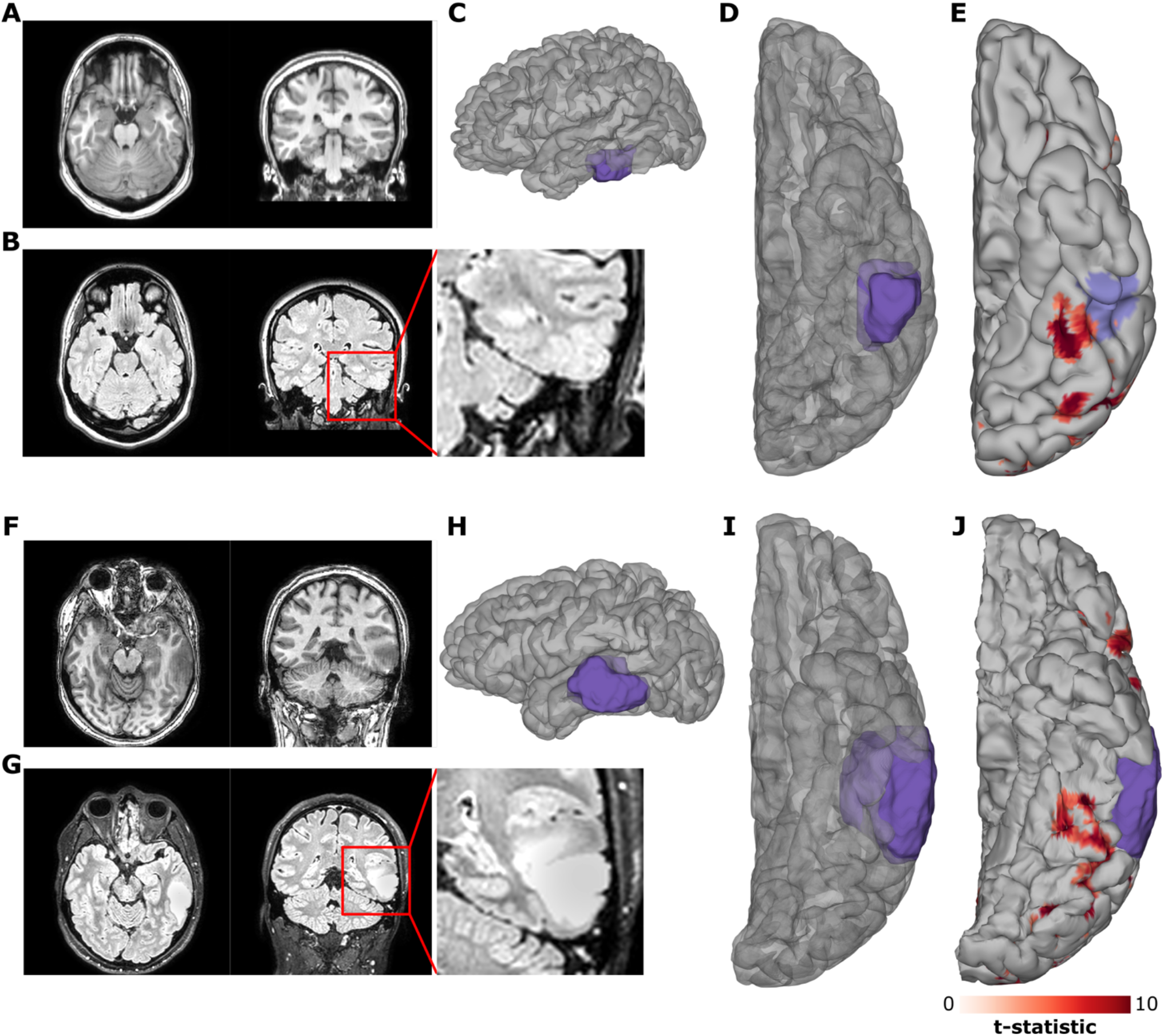
Preoperative Imaging. A,B,F,G: Preoperative T1 (A,F) and FLAIR (B,G) MRI imaging for P1 (A,B) and P2 (F,G). C,D,H,I: Lateral (C,H) and ventral (D,I) views of 3D reconstructions of the patients’ cortical surfaces and the focal cortical dysplasia (P1; C,D) and tumor (P2; H,I), as determined from the preoperative MRI. E,J: fMRI contrast of words > fixation for P1 (E) and P2 (J) with the predicted pial involvement of the relevant pathology highlighted. fMRI thresholded at t > 5.

<u>Patient 2 (P2)</u> was a right-handed male in his thirties presenting with new onset seizures resulting in aphasia. His MRI revealed a left temporal lobe tumor spanning across the inferior, middle and superior temporal gyri (Figure 2F-I).

Left hemispheric dominance of language function was confirmed with fMRI in both patients and both had word responsive fMRI clusters in close proximity to their pathological tissue (Figure 2E,J).

Multiple electrodes demonstrated strong BGA selectivity for either words or faces (Figure 3A,B). Contrasts of words and faces demonstrated spatially contiguous clusters of strong selectivity in vOTC (Figure 3C,D). In both patients, functional BGA responses were observed around the borders of their pathological tissue, as defined by the MRI. Signal quality was comparable to that seen in the extraoperative cases and there were no notable differences in the quality of neural responses between the <7 (P1) and <3 (P2) minute versions of the paradigm.

**Figure 3:**
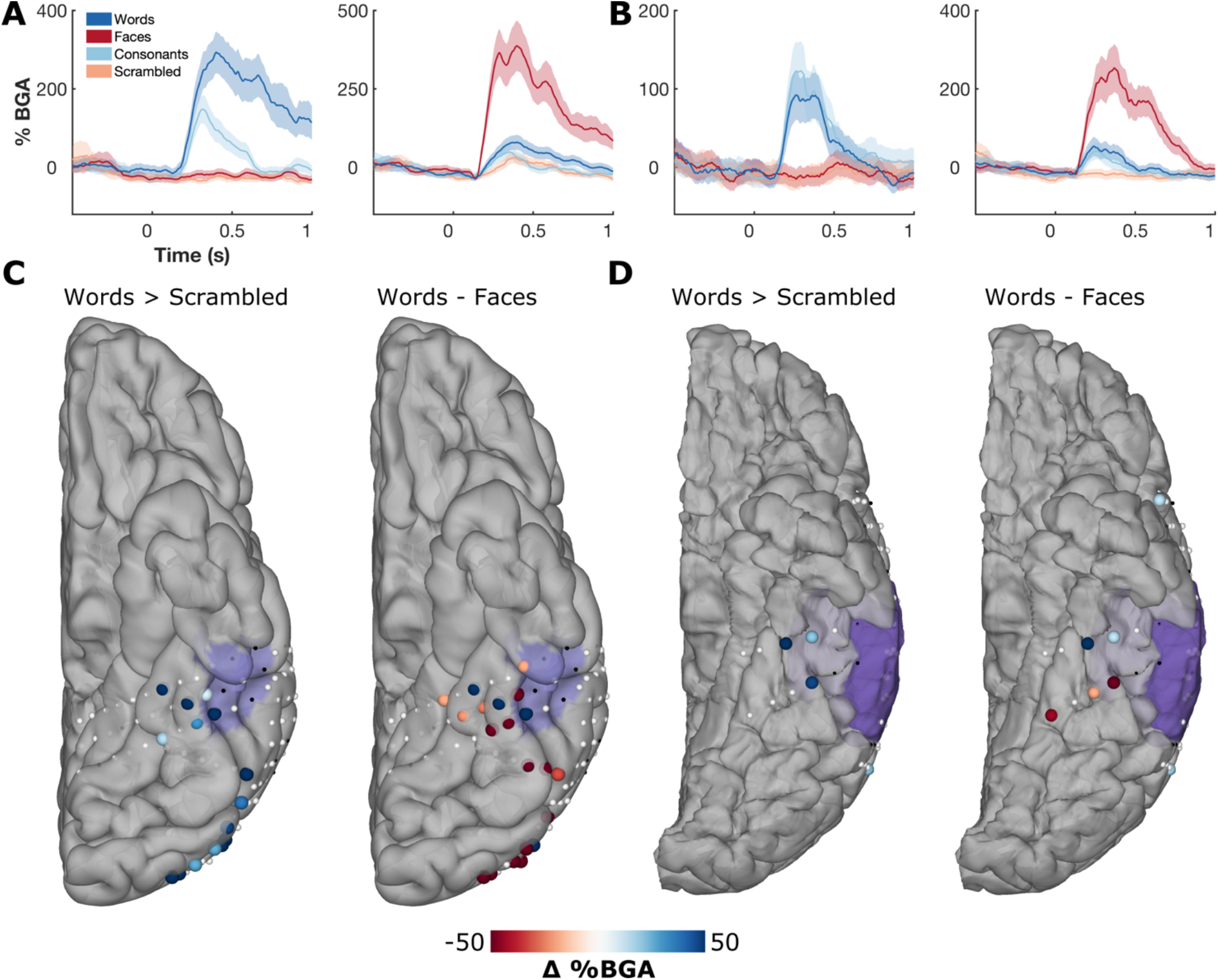
ECoG Category Selectivity. A,B: Example responses of word and face selective sites from P1 (A) and P2 (B). C,D: Spatial map of contrasts of words > scrambled, and words – faces in the window 100-400 ms after stimulus onset for P1 (C) and P2 (D). Thresholded by electrodes with a significant difference (two-sample t-test, p<0.01). Electrodes in black were excluded due to excessive inter-ictal spikes. Preoperative MRI-derived mask of pathological tissue is highlighted in purple.

### Cortical Stimulation Mapping

In Patient 1, 10 distinct sites across lateral inferior temporal gyrus and over the FCD were tested using a handheld stimulator. All of these sites were CSM and ECoG negative.

In Patient 2, the cortical patch that showed greater BGA to words than faces was stimulated using a handheld stimulator leading to sustained reading disruption (Figure 4B). The patient’s subjective reports during reading disruptions were comparable to those from previous studies of stimulation in vOTC^9^: “I see the words but they’re not coming out of my mouth”.

**Figure 4:**
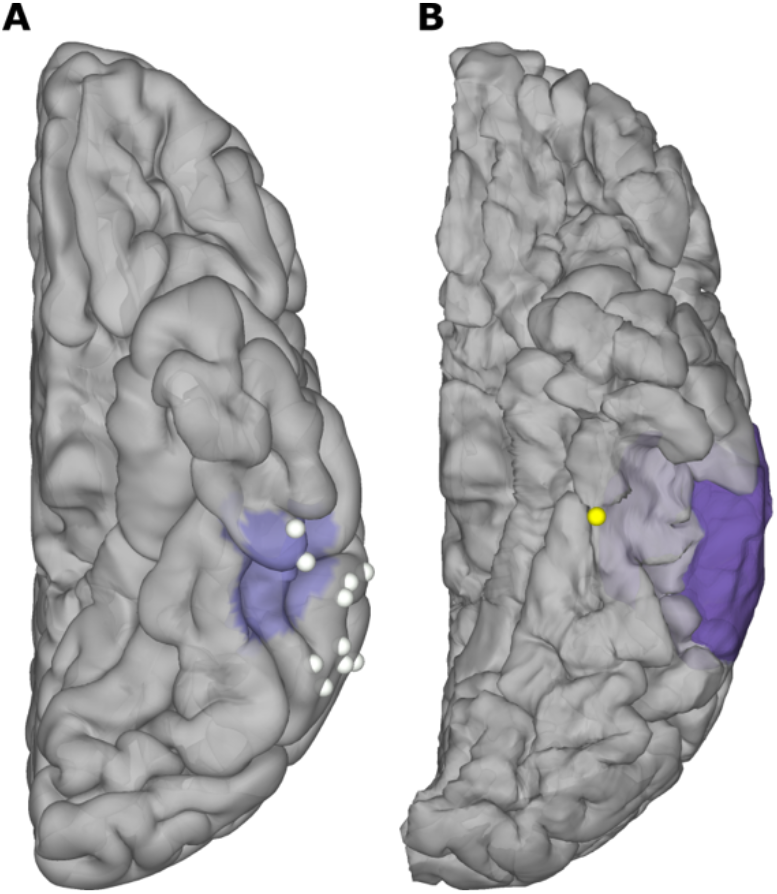
Intraoperative Stimulation. Stimulation testing during reading in P1 (A) and P2 (B), highlighting CSM positive (yellow) and negative (white) sites. MRI-derived mask of pathological tissue is highlighted in purple.

During the resection both patients were periodically tested, without stimulation, using the same behavioral tasks and displayed no noticeable deficits at any point.

## Discussion

Prior studies investigating the correspondence of task-based ECoG with cortical stimulation have primarily focused on higher-level language networks underlying visual naming across the lateral cortical surface.^17–19^ Here we show that task-based ECoG is also of utility for intraoperative mapping of ventral visual cortex, which contains several critical language sites that are mostly underappreciated in the clinical literature. Here we present evidence that intraoperative ECoG recordings in vOTC are robust, reliable and repeatable. This stimulus-induced activity can act as a rapid method of mapping function in vOTC with little active patient participation required, which further reduces the risk of functional deficits following surgery. While direct cortical stimulation is relatively time consuming, its combination with task-based ECoG increases efficiency, reduces the need for active patient participation, and provides an additional tool to localize eloquent cortex.

In the extraoperative cohort we observed a subset of sites in ventral anterior temporal lobe that resulted in stimulation-induced reading disruption but no significant ECoG activity. These sites were all anterior to regions associated with long-term reading or naming deficits as predicted by lesion studies,^29^ and may be more reflective of spreading effects caused by stimulation within pathological tissue. Additionally, we also observed that stimulation through the ECoG electrodes appeared to disrupt a broader area of cortex than is highlighted by ECoG activations. There are several likely explanations for this including that: (i) ECoG activation was derived for individual electrodes however stimulation was performed on neighboring bipolar pairs. Individual electrodes can be isolated using bipolar stimulation if they result in disruption as part of multiple tested pairs^18^; however, given time constraints, it was not possible to stimulate all electrode pairs in these cases, and testing consisted primarily of non-overlapping electrode pairs. (ii) The effects of stimulation likely affect broader areas of cortex than the ECoG signals are recorded from.^30^ It is well known that, at high currents, stimulation can cause non-local effects in functionally connected regions.^12^

## Conclusions

Given that ECoG activations are correlational rather than causal they do not act as a replacement for traditional stimulation methods. However, this method can act as an effective broad mapping tool to prioritize high probability functional regions and can serve as an additional tool when mapping eloquent cortex. While many patients would benefit from mapping techniques that require less patient participation, this mostly passive method would be of particular utility in situations where an extended stimulation mapping session is impractical, such as in patients experiencing delirium, agitation or confusion while emerging from sedation, or within pediatric populations, who often have difficulty participating in long, repetitive and highly structured tasks.^16,17^ Additionally, this method could be utilized in patients with seizure onset zones located in close proximity to functional tissue as stimulation-induced disruptions may reflect the induction of a localized seizure rather than direct disruption of eloquent cortex. Altogether our results support the conclusion that the confluence of ECoG activation and stimulation-induced deficits acts as a localizer of the VWFA in mid-fusiform cortex.^9^ As such, the use of ECoG and cortical stimulation in combination to localize brain regions essential for reading in both extraoperative and intraoperative settings is a useful approach to identify functional tissue in order to minimize postoperative reading deficits following surgical resection.^10,11^

## Data Availability

All data produced in the present study are available upon request to the authors

## Acknowledgements

We express our gratitude to all the patients who participated in this study; the neurologists at the Texas Comprehensive Epilepsy Program who participated in the care of these patients; and the nurses and technicians in the Epilepsy Monitoring Unit at Memorial Hermann Hospital who helped make this research possible.

## Abbreviations

ECoG: electrocorticography
vOTC: ventral occipitotemporal cortex
VWFA: visual word form area
CSM: cortical stimulation mapping
FCD: focal cortical dysplasia

